# Elevated 11-oxygenated androgens are not a major contributor to HPG-axis disturbances in adults with congenital adrenal hyperplasia due to 21-hydroxylase deficiency

**DOI:** 10.1101/2021.01.13.21249718

**Authors:** Matthias K. Auer, Luisa Paizoni, Meike Neuner, Christian Lottspeich, Heinrich Schmidt, Martin Bidlingmaier, James Hawley, Brian Keevil, Nicole Reisch

## Abstract

**Context:** Hypothalamus-pituitary-gonadal (HPG)-axis disturbances are a common phenomenon in patients with classic congenital adrenal hyperplasia (CAH). 11-oxygenated androgens have been suggested to play a role in this context.

**Design:** Cross-sectional single center study including 89 patients (N=42 men, N=55 women) with classic CAH.

**Main Outcome Measures:** Independent predictors for hypogonadism in men and secondary amenorrhea in women with CAH with a special focus on 11-ketotestosterone (11KT) and 11β-hydroxyandrostenedione (11OHA4).

**Results:** Hypogonadotropic hypogonadism was present in 23% of men and 61% of those women currently not on contraceptives suffered from irregular menstrual cycles or amenorrhea. Testicular adrenal rest tumor (TART) was documented in 28% of men. While 11KT (3.5x) and 11OHA4 (5.7x) among other adrenal steroids were significantly elevated in men with hypogonadism, in stepwise logistic regression, the only significant independent predictor for hypogonadism were elevated 17-OHP levels (B = 0.006; p = 0.039). Although 11KT (5.2x) and 11OHA4 (3.7x) levels were also significantly higher in women with amenorrhea in comparison to those with a regular cycle, the only significant predictor for amenorrhea were elevated total testosterone levels (B = 1.806; p = 0.040). 11-oxygenated androgens were not different in those with TART and those without. Of note, there were no significant differences in 11OHA4 or 11KT between those with a regular cycle and those currently on hormonal contraceptives.

**Conclusions:** 11-oxygenated androgens do not seem to add additional information for explaining menstrual disturbances and hypogonadism in patients with CAH in comparison to established marker of disease control.

**Highlights:**

- The 11-oxygenated androgens 11-ketotestosterone and 11β-hydroxyandrostenedione are significantly elevated in men with CAH with hypogonadotropic hypogonadism and women with amenorrhea.
- Elevated 17-OHP levels in male patients and elevated testosterone levels in female patients are however the dominant predictor of HPG-axis disturbances.
- 11-oxygenated androgens are not predictive for testicular adrenal rest tumors in men with CAH
- 11-ketotestosterone and 11β-hydroxyandrostenedione levels do not seem to be influenced by intake of oral contraceptives.

## 1. Introduction

Congenital adrenal hyperplasia (CAH) encompasses a group of hereditary autosomal recessive disorders that are characterized by mutations in key enzymes of cortisol synthesis. Among these, patients affected by mutations in the 21-alpha hydroxylase gene are the largest entity (1). Depending on the severity of the underlying mutation, patients are either suffering from isolated glucocorticoid-(simply virilizing form= SV) or additional mineralocorticoid-deficiency, when less than 1-2% of enzymatic activity is retained (salt-wasting = SW). Glucocorticoid deficiency in turn results in ACTH overstimulation due to the loss of hypothalamo-pituitary negative feedback which subsequently drives excessive adrenal androgen production (2).

Hypothalamus-pituitary-gonadal (HPG)-axis disturbances are a common phenomenon in CAH patients of both sexes. In particular in women in the fertile age, menstrual disturbances occur in up to 30% (3-5). The most obvious mechanism for this finding is the inherent extra-gonadal androgen and progesterone production in CAH. Correspondingly, menstrual cycle disturbances in female patients with CAH have been associated with poor treatment control as indicated by increased 17-OHP, testosterone, progesterone and androstenedione levels (6). In line, a recent study from the DSD-life cohort demonstrated that an androstenedione/testosterone ratio ≥1, indicating excessive adrenal testosterone production, is a strong predictor of sub-normal testosterone levels in men with CAH (7). Most of these patients correspondingly present with low or inadequately low gonadotropin levels indicating hypogonadotropic hypogonadism, while some patients also show signs of primary testicular damage mainly due to testicular adrenal rest tumor (TART) formation (7-9)

However, HPG-axis disturbances can also be found in patients who are regarded well controlled based on the aforementioned treatment markers (4,10). The exact culprit for HPG disturbances is therefore not always clear, as all major steroids, namely testosterone, estradiol and progesterone are important parts of the HPG feedback loop (11).

In recent years, a further class of adrenal-derived androgens has come into focus in the context of CAH. These are 11-oxygenated 19-carbon androgens, in particular 11-ketotestosterone (11KT) and 11β-hydroxyandrostenedione (11OHA4). Androstenedione can be converted to 11OHA4 by 11β-hydroxylase, then to 11-ketoandrostenedione through the action of 11β-hydroxysteroid dehydrogenase type 2 and finally to 11KT (12,13). It has been demonstrated that 11KT and its derivate 11KT-dihydrotestosterone act at the androgen receptor with equal potency to the classic testosterone and dihydrotestosterone (DHT) (14). Correspondingly, it has been shown that these steroids are significantly elevated in patients with classic CAH (15). While it has been suggested that elevated 11KT levels might contribute to HPG-suppression at the hypothalamic and pituitary level, their clinical relevance has still to be determined (16). Although testosterone exerts direct negative effects at the hypothalamic levels via AR binding (17,18), estradiol seems to be the dominant suppressor in this context (19). As 11KT cannot be directly aromatized questions a dominant role in HPG suppression in this context (14).

In the present study, we were therefore interested if 11-oxygenated androgens, in particular 11-KT would add information on HPG-axis disturbances in adult patients with CAH due to 21-hydroxylase deficiency in addition to established markers of disease control.

In detail, we hypothesized that 11KT would be higher in men with hypogonadotropic hypogonadism than in those with eugonadism, that 11OHA4 would be higher in males with TART than in those without and that 11KT and 11OHA4 would be higher in women with menstrual irregularities in contrast to those with eumenorrhoea. In an exploratory analysis we were further interested if 11KT and 11OHA4 would differ between women with a regular cycle and those on oral contraceptive therapy to estimate if these potential covariates should be accounted for in the clinical context if our main hypotheses would hold true.

## 2. Material and Methods

### Subjects

We included 89 patients (N=42 men, N=55 women) of whom 58 were suffering from SW and 31 of the SV form of CAH. All patients were seen at the outpatient clinic of the Medizinische Klinik IV, Klinikum der Universität München, LMU Munich. All patients were directly approached during a regular visit in the outpatient clinic. The recruitment period was from January 2012 to January 2014. The local ethics committee of the Ludwig-Maximilians-Universität München approved the study protocol. All participants provided their written informed consent.

### Testicular ultrasound

Testicular ultrasound was performed in the Department of Endocrinology in the University Hospital Munich by a trained sonographer (Siemens Acuson S2000). Testicular ultrasound was performed documenting testicular masses. Greyscale and color Doppler ultrasonography were obtained in the longitudinal and transverse planes using a 15MHz transducer. Data on TART was missing in seven men.

### Hormone assays and hormonal control

Blood was drawn in fasting patients in the morning between 8 and 10 a.m., roughly 2 hours after the intake of the morning medication of participants. Concentrations of adrenocorticotrophic hormone (ACTH), sex hormone binding globulin (SHBG) and DHEAS were determined using the Liaison chemiluminescence immunoassays (DiaSorin, Sallugia, Italy).

Total testosterone, 17-OHP, androstenedione, 11-KT and 11OHA4 were quantified by LC-MS/MS. Steroid extraction and quantitation was carried out as previously described (20).

Daily doses of glucocorticoids and mineralocorticoids were evaluated. Equal hydrocortisone doses were calculated according to their suggested androgen-suppressive capacity (hydrocortisone=1, prednisolone=4, dexamethasone=70) and adjusted to the patients’ body surface area (2). In men, hypogonadism was defined as having a low testosterone level <12nmol/l (21)(39). Hypogonadotropic hypogonadism was defined as having low testosterone levels with inappropriately normal or low LH/FSH. Secondary amenorrhea was defined as not having had a spontaneous menstrual bleeding during the last six months prior to study inclusion, while irregular cycles were defined as having as cycle length that was less than 21 or more than 35 days (22).

### Statistical analysis

Statistical analysis was performed using PASW Statistics (formerly SPSS) Version 22.0 for Windows. Sample characteristics were compared using χ2 tests for categorical variables and the Mann-Whitney U test for continuous variables.

Spearman correlation was performed for univariate correlations. Partial regression analyses with LH and FSH as a dependent variable and potential influential factors as independent variables were performed adjusted for age and BMI. Separate multivariate regression analyses were used to identify independent contributors for hypogonadism in men and secondary amenorrhea in women. Due to the strong correlation among most steroid measures, variables were mean centered before addition to the model. A two-block linear regression analysis was carried out to assess the contribution of different variables. Block 1 included age and BMI using a forced entry method. In Block 2, we added CAH-specific covariates using forward stepwise regression analysis. A two-sided P value of <0.05 was considered statistically significant.

## 3. Results

### General characteristics

The median age of the cohort was 28 years (range 18-61). The median GC equivalence dosage was 16.0 mg/m2/d for men and 14.6 mg/m2/d for women. According to the expected (residual) enzymatic activity, thirteen (21.0% of whom data was available) patients were assigned to genotype group 0, 21 (33.9%) to genotype group A, 24 (38.7%) to genotype group B and four (6.5%) patients to genotype group C (23).

Of those investigated, TART was present in eleven patients (28.2%). 23% presented with hypogonadotropic hypogonadism. There were no men with primary hypogonadism.

Thirty seven percent of women were currently receiving hormonal contraceptive therapy. From those without oral contraception and in premenopausal age, 38.7% had a regular cycle while 25.8% had irregular menses and 35.5% were amenorrhoeic. Three women were postmenopausal (6%). At study inclusion, of those with a regular cycle, 75% were in the follicular and 25% in the luteal phase of the cycle (Table 1).

**Table 1.** General characteristics.

|  |  | Men |  | Women |  |  |  |
| --- | --- | --- | --- | --- | --- | --- | --- |
|  |  | N (39) | % | N (50) | % |  | p |
| Mutation |  |  |  |  |  |  |  |
| Mutation group | 0 | 8 | 21 | 5 | 10 |  | 0.293 |
|  | A | 12 | 31 | 9 | 18 |  |  |
|  | B | 8 | 21 | 16 | 32 |  |  |
|  | C | 2 | 5 | 2 | 4 |  |  |
|  | Unknown | 9 | 23 | 18 | 36 |  |  |
| Simply virilizing |  | 12 | 31 | 19 | 38 |  |  |
| Salt wasting |  | 27 | 69 | 32 | 64 |  | 0.477 |
| TART | Yes | 11 | 28.2 | NA | NA |  |  |
|  | unilateral | 3 |  | NA | NA |  |  |
|  | bilateral | 7 |  | NA | NA |  |  |
|  | No | 21 | 53.8 | NA | NA |  |  |
|  | Missing | 7 | 17.9 | NA | NA |  |  |
| Hypogonadism |  |  | 0 |  |  |  | NA |
|  | Eugonadism | 30 | 77 | NA | NA |  |  |
|  | Sec. hypogonadismus | 9 | 23 | NA | NA |  |  |
|  | Primary hypogonadism | 0 | 0 | NA | NA |  |  |
| Cycle | Oral contraception | NA | NA | 16 | 32 |  | NA |
|  | Regular cycle | NA | NA | 12 | 24 | 38.7* |  |
|  | Follicular | NA | NA | 9 | 12 | 75%** |  |
|  | Luteal | NA | NA | 3 | 8 | 25%** |  |
|  | Irregular menses | NA | NA | 8 | 16 | 25.8* |  |
|  | Secondary amenorrhea | NA | NA | 11 | 22 | 35.5* |  |
|  | Postmenopausal | NA | NA | 3 | 6 |  |  |
|  |  |  | 0 |  | 0 |  |  |
| GC | None | 0 | 0 | 2 | 4 |  | 0.359 |
|  | HC | 18 | 46 | 19 | 38 |  |  |
|  | Predniso-ne/-lone | 16 | 41 | 24 | 48 |  |  |
|  | Dexa | 4 | 10 | 3 | 6 |  |  |
|  | HC + Dexa | 0 | 0 | 3 | 6 |  |  |
|  | Predniso-ne/-lone +Dexa | 1 | 3 | 0 | 0 |  |  |
|  |  | **Median** |  | **Median** |  |  |  |
| HC Equivalence-dosage (mg/m<sup>2</sup>) |  | 14.6 |  | 16.0 |  |  | 0.143 |

*Man-Whitney U, X2 test*
*\*Of those not on contraception*
*\*\*of those with regular cycle*
*NA not applicable*
*TART: testicular adrenal rest tumor*
*HC: hydrocortisone*
*GC: glucocorticoid*
*Dexa: Dexamethasone*
*TART: Testicular adrenal rest tumor*

There were no significant differences in androstenedione,17-OHP,11KT and 11OHA4 levels between men and women. (Table 2).

**Table 2.** Anthropometric measures and hormone values.

|  | Men |  |  | Women |  |  |  |
| --- | --- | --- | --- | --- | --- | --- | --- |
|  | Median | Percentiles |  | Median | Percentiles |  | p |
|  |  | 25 | 75 |  | 25 | 75 |  |
| Age | 28.0 | 24.0 | 40.0 | 29.0 | 24.0 | 38.0 | 0.990 |
| BMI (kg/m²) | 26.0 | 22.7 | 29.9 | 26.8 | 22.3 | 30.8 | ND |
| Waist (cm) | 95.0 | 81.3 | 100.8 | 78.0 | 70.0 | 92.0 | ND |
| Body fat (%) | 25.6 | 13.1 | 25.7 | 34.3 | 18.5 | 33.2 | ND |
| Lean mass (%) | 57.1 | 52.5 | 63.1 | 44.4 | 40.1 | 48.8 | ND |
| LH (U/l) | 3.0 | 1.8 | 4.7 | 3.4 | 0.9 | 5.7 | ND |
| FSH (U/l) | 5.0 | 2.8 | 7.2 | 4.6 | 2.8 | 6.9 | ND |
| ACTH (pg/ml) | 11.0 | 6.0 | 55.0 | 10.0 | 4.0 | 29.0 | 0.219 |
| Testosterone (nmol/l) | 13.0 | 10.9 | 16.7 | 0.5 | 0.2 | 1.5 | ND |
| Androstenedione (nmol/l) | 3.6 | 1.1 | 8.2 | 2.0 | 0.9 | 6.0 | 0.155 |
| 17-OHP (nmol/l) | 23.3 | 5.2 | 153.6 | 14.0 | 2.8 | 64.5 | 0.125 |
| 11KT (nmol/l) | 1.2 | 0.3 | 2.4 | 0.6 | 0.2 | 2.4 | 0.335 |
| 11OHA4 (nmol/l) | 2.0 | 0.3 | 8.0 | 1.3 | 0.3 | 5.5 | 0.550 |
| DHEAS (µg/ml) | 0.3 | 0.2 | 0.5 | 0.0 | 0.0 | 0.2 | ND |
| SHBG (nmol/l) | 33.5 | 24.1 | 39.1 | 57.8 | 39.2 | 145.1 | ND |
| A/T-ratio | 0.30 | 0.1 | 0.7 | 3.7 | 3.5 | 4.8 | ND |
| 11KT/T-ratio | 0.1 | 0.02 | 0.24 | 1.2 | 1.0 | 1.7 | ND |
| FAI (%) | 12.4 | 9.0 | 17.2 | 0.2 | 0.1 | 0.8 | ND |
| HOMA-IR | 1.6 | 1.1 | 2.0 | 2.2 | 1.0 | 3.4 | 0.190 |

258 *Mann-Whitney U*259 *ND not determined*260 *FAI: free androgen index*261 *11OHA4: 11-β hydroxyandrostenedione*262 *11KT: 11-ketotestosterone*263 *T: Testosterone*

### Correlations between steroid measures

In men, there was a strong positive correlation between 11KT with ACTH (0.845; p < 0.001), androstenedione (R = 0.933; p < 0.001), 17-OHP (R = 0.942; p < 0.001) and the androstenedione/testosterone (A/T)-ratio (R = 0.837; p < 0.001). A negative correlation of 11KT with testosterone was only seen on a trend level (R = -0.278; p = 0.087) and there was no significant correlation with the free androgen index (FAI). 11OHA4 correlated positively with ACTH (R = 0.868; p < 0.001), androstenedione (0.944; p < 0.001), 17-OHP (R = 0.947; p < 0.001) and the A/T-ratio (R = 0.857; p < 0.001). 11KT and 11OHA4 also were strongly correlated (R = 0.976; p< 0.001).

In women, there was a positive correlation of 11KT with ACTH (R = 0.552; p < 0.001), total testosterone (R = 0.899; p < 0.001), androstenedione (R = 0.889; p< 0.001), 17-OHP (R = 0.915; p < 0.001), DHEAS (R = 0.618; p < 0.001) and the FAI (R = 0.780; p < 0.001). 11OHA4 correlated positively with ACTH (R = 0.604; p < 0.001), total testosterone (R = 0.875; p < 0.001), androstenedione (R = 0.886; p< 0.001), 17-OHP (R = 0.915; p < 0.001), DHEAS (R = 0.637; p < 0.001) and the FAI (R = 0.781; p < 0.001). The correlation between 11KT and 11OHA4 was weaker than in men (R = 0.477; p< 0.001).

### Presence of TART

There were no significant differences in steroid measures between those with and without TART. There was also no difference in ACTH or gonadotropin levels, HC equivalence dosage or SV vs. SW (data not shown).

### Hypogonadism in men

Androstenedione levels were significantly higher in men with secondary hypogonadism than in those with eugonadism (4.5x; p = 0.023). A similar pattern could be observed with 17-OHP (14.4x; p = 0.007), 11KT (3.5x; p = 0.020), 11OHA4 (5.7x, p = 0.009) and ACTH (9.3x; p = 0.004) (Figure 1).

**Figure 1.**
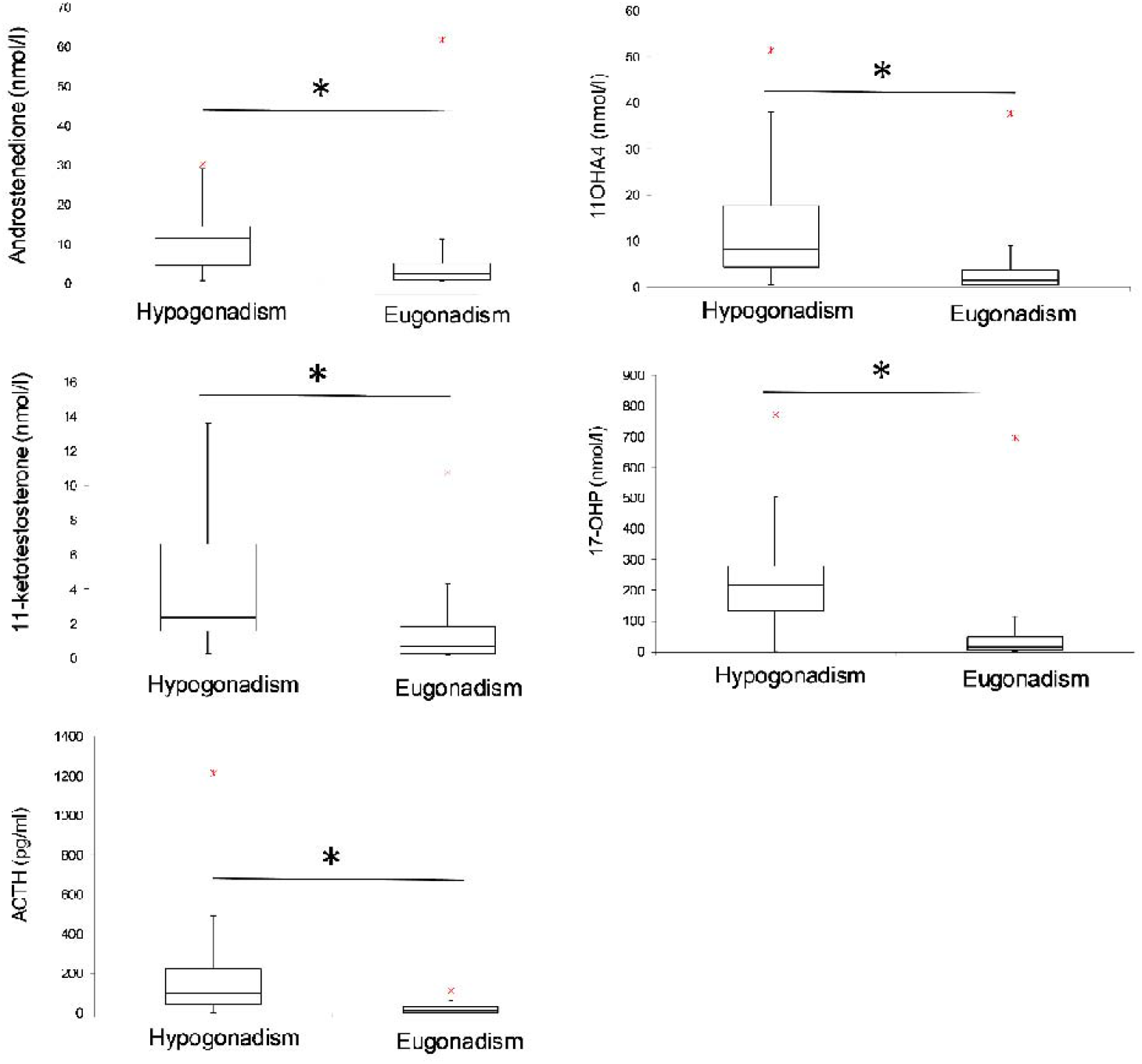
Differences between men with hypogonadism and eugonadism. Androstenedione levels were significantly higher in men with secondary hypogonadism than in those with eugonadism (4.5x; p = 0.023). Same was true for 17-OHP (14.4x; p = 0.007), 11KT (3.5x; p = 0.020), 11OHA4 (5.7x, p = 0.009) and ACTH (9.3x; p = 0.004). (Boxes represent the first to the third quartile, vertical line indicates medians, crosses indicate outliers).

### LH/FSH in men

In men, according to partial correlation analysis, adjusted for BMI and age, there was no significant correlation of LH with any steroid measure. There was also no correlation with HC equivalence dosage, type of GC (HC = 0 vs. synthetic GC = 1) or with regard to SV vs. SW.

In contrast, there were comparably strong negative correlations of androstenedione (R = -0.330; p = 0.048), 17-OHP (R = -0.329; p = 0.047); 11KT (R = -0.368; p = 0.025), 11OHA4 (R = -0.361; p = 0.028), the A/T-ratio (R = -0.387; p = 0.018) as well as the KT/T-ratio (R = -0.383; p = 0.019) with FSH. There was no significant correlation with LH or FSH when adding KT+T as a new variable, given then idea that both androgens should have equimolar androgenic potency according to the literature.

### Predictors of hypogonadism in men

The only significant predictor for hypogonadism in men were elevated 17-OHP levels (B = 0.006; p = 0.039) in a stepwise logistic regression (0 = Eugonadism, 1= hypogonadism), including age, BMI, ACTH, androstenedione, 17-OHP, DHEAS, 11KT and 11OHA4. As expected, finding also remained significant when adding HC-equivalence dosage as well as HC (0) vs GC (1) to the model (Table 3).

**Table 3.**
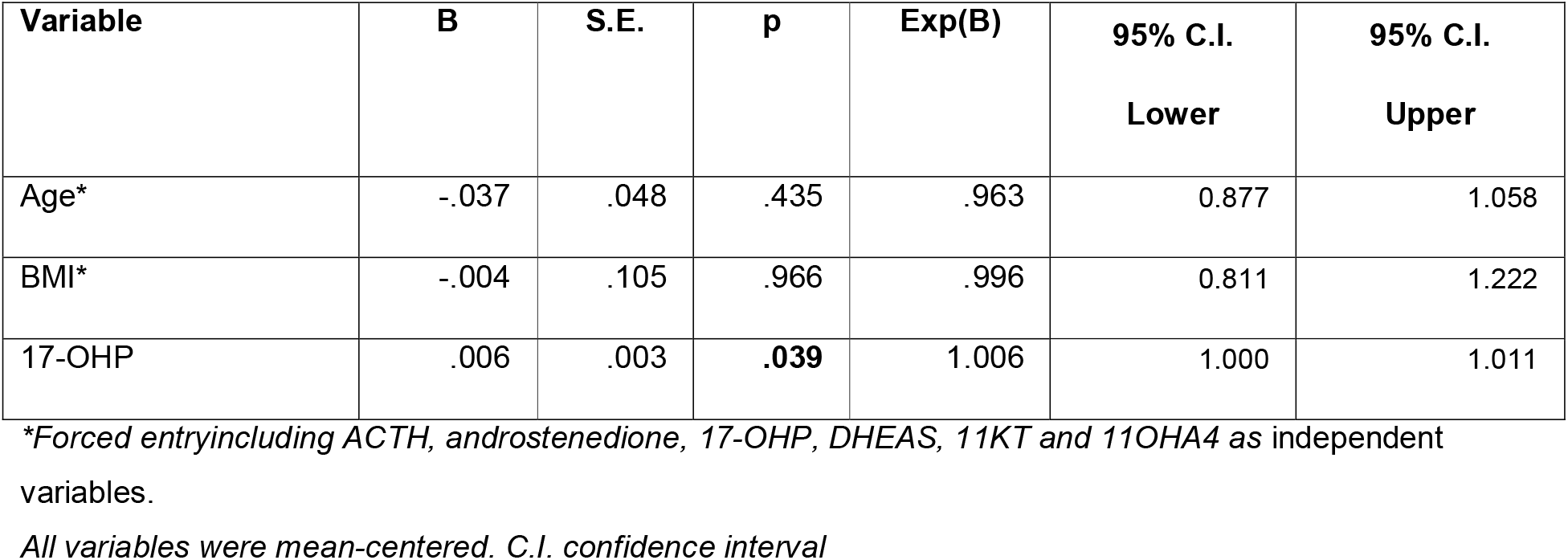
Stepwise regression of potential predictors of hypogonadism in men.

### Amenorrhea in women

ACTH (4.3x; p = 0.003), androstenedione (2.5x; p = 0.019), 17-OHP (8.1x; p = 0.002), 11KT (5.2x; p = 0.002), 11OHA4 (3.7x; p = 0.023) and testosterone (3.1x; p = 0.007) were significantly higher in women with amenorrhea versus regular cycling women (Figure 2).

**Figure 2.**
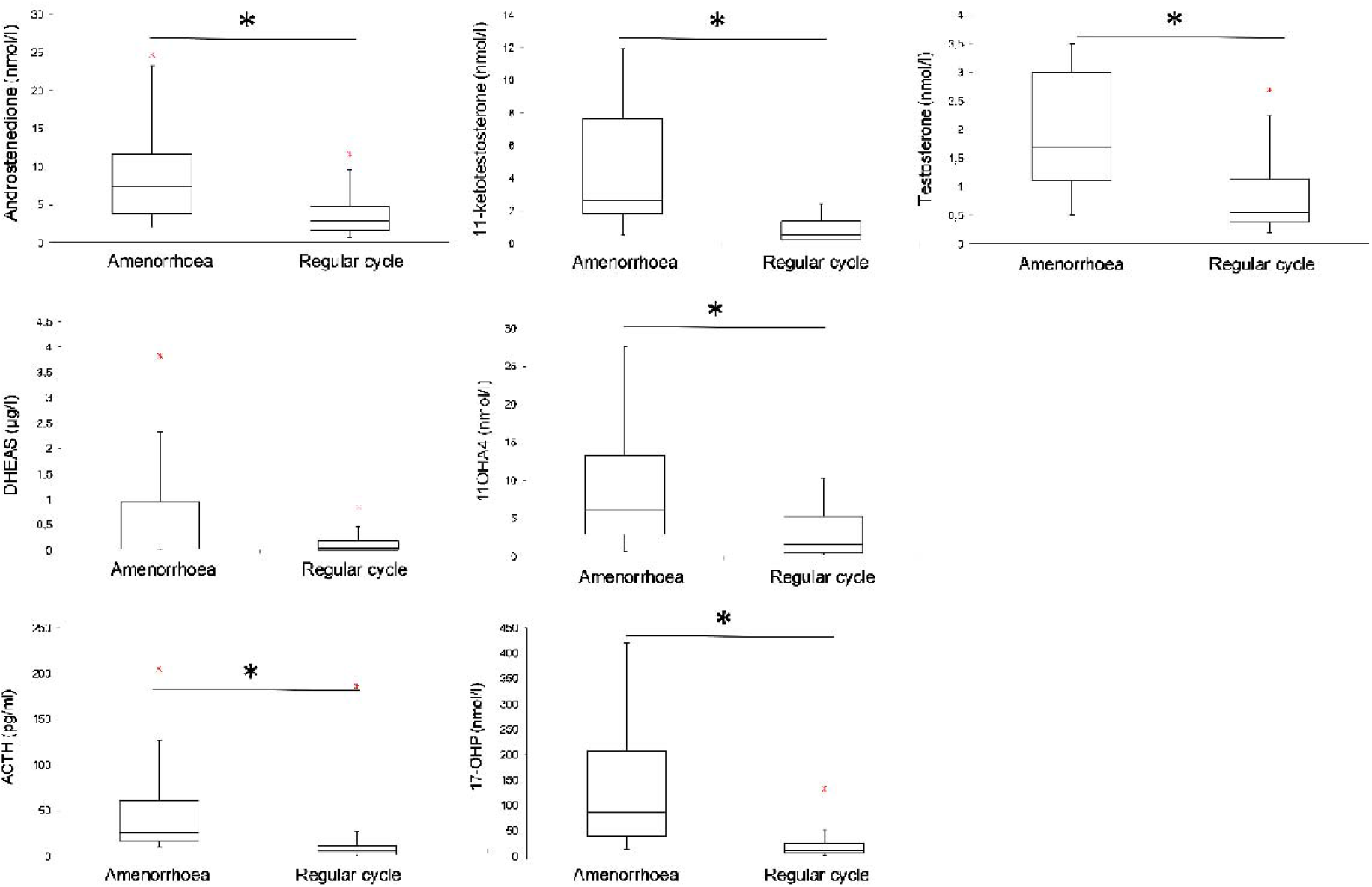
Differences between regular cycling and amenorrhoeic women. ACTH (4.3x; p = 0.003), androstenedione (2.5x; p = 0.019), 17-OHP (8.1x; p = 0.002), 11KT (5.2x; p = 0.002), 11OHA4 (3.7x; p = 0.023) and testosterone (3.1x; p = 0.007) were significantly higher in women with amenorrhea in comparison to those with a regular cycle. (Boxes represent the first to the third quartile, vertical line indicates medians, crosses indicate outliers).

Equivalence dosage of HC was lower (12.7mg/m^2^ vs 16.4mg/m^2;^ p = 0.019). There was no difference in terms of GC vs HC use or HOMA-IR. There was also no difference in DHEAS levels and the A/T-ratio. There was no significant difference in these parameters between those with irregular and those with a regular cycle (data not shown).

### Oral contraceptive use

Androstenedione (p = 0.011), the FAI (p = 0.001) and DHEAS (p = 0.049) were significantly lower in women with a regular menstrual cycle in comparison to women currently taking an oral contraceptive pill, while there was no difference in total testosterone, 17-OHP, 11KT or 11OHA4 (Figure 3).

**Figure 3.**
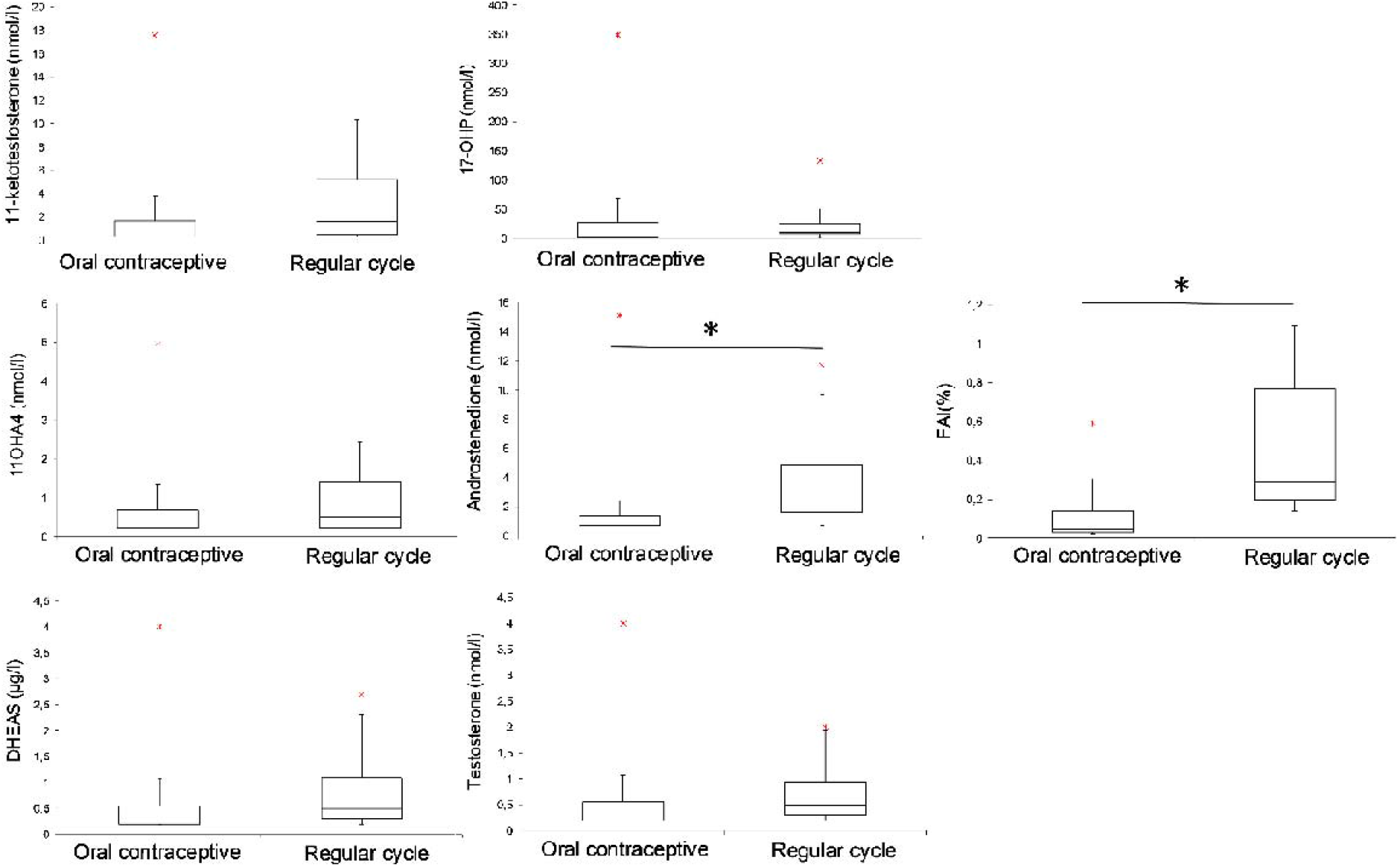
Oral contraception vs regular cycle. When comparing those with a regular menstrual cycle with those currently taking an oral contraceptive pill, androstenedione (p = 0.011), the FAI (p = 0.001) and DHEAS (p = 0.049) were significantly lower, while there was no difference in total testosterone, 17-OHP, 11KT or 11OHA4.). (Boxes represent the first to the third quartile, vertical line indicates medians, crosses indicate outliers).

### Predictors of amenorrhea

Only higher testosterone levels were significantly associated with amenorrhea (B = 1.806; p = 0.040) in a logistic regression analysis including age, BMI (Enter) (0 = amenorrhea, 1= regular cycle), ACTH, androstenedione, 17-OHP, DHEAS, total testosterone, free testosterone, the A/T-ratio, 11KT, 11OHA4, and the T/KT-ratio (stepwise) (Table 4).

**Table 4.** Stepwise regression of potential predictors of amenorrhea.

| Variable | B | S.E. | p | Exp(B) | 95% C.I. | 95% C.I. |
| --- | --- | --- | --- | --- | --- | --- |
|  |  |  |  |  | Lower | Upper |
| Age* | -0.097 | 0.111 | 0.381 | .908 | 0.731 | 1.127 |
| BMI* | -0.024 | 0.095 | 0.804 | .977 | 0.812 | 1.176 |
| Testosterone | 1.806 | 0.877 | **0.040** | 6.085 | 1.091 | 33.955 |
| Constant | 0.796 | 0.803 | 0.321 | 2.216 |  |  |

\*Forced entry; Stepwise including ACTH, androstenedione, 17-OHP, DHEAS, total testosterone, free testosterone, the A/T-ratio, 11KT, 11OHA4, and the T/KT-ratio as independent variables. All variables were mean-centered. C.I. confidence interval.

## 4. Discussion

We could demonstrate in this relatively large cohort that 11-oxygenated androgens are significantly elevated in patients of both sexes with HPG-axis disturbances. They however do not seem to be the most relevant androgens in this context.

The prevalence of hypogonadism in men and menstrual irregularities in women in our cohort was comparable to that reported in the literature (7,24) and hence emphasizes that HPG-axis disturbances are still a fairly common issue in this patient population. Despite a significant elevation of 11KT and 11OHA4, being in accordance with earlier studies (16), 17-OHP was a better predictor of hypogonadism in male patients. Our data therefore do not indicate a leading role of 11KT in HPG-axis suppression. This may be due to the fact that 11KT cannot be converted into estrogenic steroids and many effects that had originally been attributed to testosterone in men are actually mediated via aromatization to estradiol (25). This also includes sex steroid regulation at the hypothalamus/pituitary level (19).

The significant role of 17OHP may be explained by the fact that progesterone is also a very potent suppressor of the HPG-axis in men (26,27). As 17-OHP is a direct metabolite of progesterone, it is a potent agonist of the PR (28) as also indicated by a strong positive correlation between both steroids in patients with CAH (29).

In contrast to previous work (16) and our initial hypothesis, neither 11KT nor 11OHA4 were elevated in men with TART and most steroid measures, including 11KT and 11OHA4 showed a negative instead of a positive correlation with FSH as a potential marker of gonadal damage. Although our cohort was age-wise comparable to those adult patients reported by Turcu et al., a considerable proportion of their patients with TART were children. While TARTs in younger patients might still respond to HPA-axis suppression, they seem to become less modifiable in the long-term due to fibrotic modifications (30). In line, presence of TART has been associated with poor treatment quality in some (8) but not all studies (9,31). Despite the factor of age, we cannot exclude that there were further differences in non-recorded potential confounders such as long-term disease control distinguishing our patient groups.

11KT and 11OHA4 levels were also higher in women with secondary amenorrhea than in those with a regular cycle, as having been reported before (16). But again, when adding other markers of disease control to our model, testosterone was superior in explaining amenorrhea. Correspondingly, menstrual cycle disturbances in female patients with CAH have been associated with poor androgen control (6). Testosterone modulates GnRH-pulse frequency and therefore interferes with the complex hormonal interplay during the menstrual cycle (32). In addition, it also has direct effects on the endometrium (33).

Of note, there were no significant differences in 11KT or 11OHA4 between those with a regular cycle and those currently taking hormonal contraceptives. This may be due to a lower binding affinity for SHBG that has been suggested but not yet demonstrated for these steroids. This could be of clinical relevance, as this might explain why some patients benefit less from oral contraceptive treatment with regard to hyperandrogenic symptoms such as hirsutism and acne (34). However, prospective studies are needed to substantiate this finding.

There are several limitations of our study. Firstly, due to the cross-sectional design we can only show associations and no causality between dependent and independent variables. Prospective studies with modifying interventions are therefore needed. A further limitation in this context is that, in contrast to the general recommendation, the definition on hypogonadism in our study was based on a single, instead of two testosterone measures in the morning (21). However, diurnal variance of testosterone and LH in CAH do generally differ from the general male population due to the extra-gonadal androgen production and also depend on GC pharmacokinetics (1). A further limitation of our study is that we only used a single steroid measurement in the morning for our analyses. Steroid measurements from blood drawn before the intake of the morning GC dosage might therefore yield other results. However, all patients included were either receiving a morning dosage of HC or prednisolone or an evening dosage of dexamethasone which show, given the wide inter-individual variability, a comparable suppression profile for the chosen time window (35,36).

Lastly, although we in investigated a large group of adult patients with classic CAH, given the rarity of the disease, subgroups were rather small, stepwise regression had to be used for our final models. Nonetheless, given the strong differences in steroid markers between groups we consider our results to be valid.

## 5. Conclusion

In the present study, 11KT and 11OHA4 did not seem to add additional information to the understanding of HPG-axis disturbances in CAH. Further studies have to show if 11oxC19 steroids are of relevance for explaining for other health issues in CAH. It might be of particular interest, if they could be a superior biomarker of disease control. To our knowledge it has not yet been shown to what extent 11-oxygenated androgens show a diurnal variation, potentially addressing the still problematic issue of timing of blood sampling with regard to different GC treatment regimens. Former approaches to overcome these restrictions like serial salivary (36) or blood sampling (37), or hair steroid measures (52) are on the one hand very time consuming and on the other hand have so far not yet been proven to be superior in terms of predicting clinically relevant long-term outcomes. Lastly, it would have to be demonstrated that 11KT can be independently targeted by treatment-modification, independent of other adrenal androgens.

## Data Availability

Data is can be provided by the corresponding author on reasonable request

## Abbreviations

11KT: 11-ketotestosterone
11OHA4: 11β-hydroxyandrostenedione
17-OHP: 17α-hydroxyprogesterone
ACTH: Adrenocorticotropic hormone
AR: Androgen receptor
CAH: Congenital adrenal hyperplasia
CPA: Cyproterone acetate
DHEAS: Dehydroepiandrosterone sulfate
ER: Estrogen receptor
FC: Fludrocortisone
FSH: Follicle-stimulating hormone
GC: Glucocorticoid(s)
GnRH: Gonadotropin releasing hormone
HC: Hydrocortisone
HPG: Hypothalamus-pituitary-gonadal
LH: Luteinizing hormone
PR: Progesterone receptor
SERM: Selective estrogen receptor modulator
SHBG: Sex hormone binding globulin
SV: Simply virilizing
SW: Salt-wasting
TART: Testicular adrenal rest tumor

## Acknowledgments

We thank all participants for taking part in this study

